# Use of an ultrasound picture archiving and communication system (PACS) to answer research questions: description of data cleaning methods

**DOI:** 10.1101/2022.12.11.22282862

**Authors:** Matthew K Moore, Gillian Whalley, Gregory T Jones, Sean Coffey

## Abstract

**Background:** Ultrasound picture archiving and communication system (PACS) databases are useful for quality improvement and clinical research, but frequently contain free text that is not easily readable. Here, we present a method to extract and clean a semi-structured echocardiography (cardiac ultrasound) PACS database.

**Methods:** Echocardiography studies between 1 January 2010 and 31 December 2018 were extracted using a data mining tool. Numeric variables were recoded with extreme values excluded. Analysis of free text, including descriptions of the heart valves and right and left ventricular size and function, was performed using a rule-based system. Different levels of free text variables were initially identified using commonly used phrases, and then iteratively developed. Randomly selected sets of 100 studies were compared to the electronic health record to validate the data cleaning process.

**Results:** The data validation step was performed three times in total, with the Cohen’s kappa ranging between 0.88 and 1.00 for the final set of data validation across all measures.

**Conclusion:** Free text cleaning of semi-structured PACS databases is possible using freely available open source software. The accuracy of this method is high, and the resulting dataset can be linked to administrative data to answer research questions. We present a method that could be used to answer clinical questions or to develop quality improvement initiatives.

## Background

Ultrasound is central to modern medical practice, with ongoing increases in clinical use seen in recent years ^1, 2^. In addition to the clinical report, local picture archiving and communication system (PACS) databases contain rich data, which can be used for quality improvement and for clinical research. However, these databases frequently contain multiple free text fields, making analysis challenging such that studies using PACS databases often only use numeric or clearly defined categorical variables.

A number of recent studies have highlighted the value of such studies. A research group in Massachusetts, United States of America has successfully examined the surveillance of mitral valve disease, by using a local database of echocardiograms with structured, granular clinical data, between 2001 and 2016 ^3^. Another team in Israel has linked their structured echocardiogram report database to nationally-collected mortality data, to examine several valvular abnormalities ^4, 5^. Work in Australia has resulted in the National Echocardiographic Database Australia (NEDA), which has already been used to look at mortality in those with moderate aortic stenosis and pulmonary hypertension ^6-8^.

However, semi-structured or unstructured PACS databases can make the conversion of qualitative variables into usable data difficult, which may discourage researchers from doing so. Frequently results from such studies are presented without providing sufficient information on the extensive data cleaning (especially of free text data) and validation processes to allow other investigators the ability to implement such processes in their local database. In this paper, we present a methodology for cleaning a semi-structured echocardiography database, and hope that the methods described here could be used by other researchers interested in examining their own ultrasound databases.

## Methods

### Ethical approval

Consultation with Māori was undertaken with the Ngāi Tahu Research Consultation Committee. This study received ethical approval from the Central Health and Disability Ethics Committee in New Zealand (ref: 21/CEN/15). Locality approval was sought from and provided by the Southern District Health Board.

### Setting and participants

The Southern District Health Board (SDHB) provides tertiary cardiology services to the lower part of the South Island, New Zealand, with approximately 5000-6500 echocardiograms performed annually for the approximately 330,000 population. The study cohort comprised all patients who had an echocardiogram at Dunedin Hospital between January 1^st^ 2010 and December 31^st^ 2018.

### Echocardiography report extraction

The echocardiographic Picture Archiving and Communication System (PACS) used in SDHB is Syngo Dynamics (version VA20F, Siemens Healthineers, Erlangen, Germany). Data was extracted using the proprietary Syngo Dynamics Data Miner, and output as a comma-separated values (csv) file. There were several hundred variables accessible to the data miner, and the vast majority of these were not in use. Individual variable names differ between different versions of the clinical echocardiography report, and so manual identification is needed for extraction. We wished to extract a set of variables that would provide the relevant information about the aortic valve, alongside key markers of heart function. For brevity, these are included in the supplementary material (supplementary table 1).

**Table 1:** Order of data cleaning. NA: not available. BMI: body mass index. IVSd: interventricular septum end-diastole. LVPWd: left ventricular posterior wall end-diastole. LVEF: left ventricular ejection fraction. LVOT: left ventricular outflow tract. AoV: aortic valve. VTI: velocity-time integral. RVSP: right ventricular systolic pressure

| Data cleaning step | Variables cleaned |
| --- | --- |
| Data loading | Data loaded into R, stitched together, and anonymized |
| Renaming | Variables renamed for ease of cleaning |
| Remove unused variables | Variables where every observation is NA were identified and excluded |
| Study date, age, sex, height, weight, and body mass index | Study date, patient age, and patient sex reformatted. Height and weight formatted, with manual correction to erroneous entries. Body mass index calculated, with a BMI $\geq 70\text{kg/m}^2$ or $\leq 14\text{ kg/m}^2$ excluded as out of plausible range. |
| Indication free text analysis | Free text analysis to identify indications |
| Cardiac rhythm and heart rate | Free text analysis to identify cardiac rhythm, formatting of heart rate (see below). Heart rate values above 200 beats per minute were excluded. |
| Left ventricle numeric variables | IVSd, LVPWd, LVEF, and LVOT diameter systole reformatted with impossible values ($\leq 0$) removed |
| Left ventricle and atrium free text variables | Free text analysis to identify left ventricular size and function, alongside left atrial size |
| Mitral valve | Mitral E, A, e', E/a, E/e' reformatted with impossible values ($\leq 0$) removed. Free text analysis to identify mitral stenosis, regurgitation, annular calcification, and prolapse. The presence of annular calcification or stenosis were then used to create a binary variable "any mitral calcification". |
| Aortic valve (AoV) | Ascending aorta diameter, aortic root diameter, AoV Vmax, AoV mean pressure gradient, AoV VTI, and AoV area reformatted with impossible |
| | values ($\leq 0$) removed. Aortic valve dimensionless index (DDI) calculated as LVOT VTI divided by AoV VTI. Free text analysis to identify AoV stenosis, AoV regurgitation, and AoV morphology/structure. |
| Pulmonary valve | Free text analysis to identify pulmonary stenosis and regurgitation. |
| Tricuspid valve | Free text analysis to identify tricuspid regurgitation. Impossible values of tricuspid regurgitation Vmax ($\leq 0$) were removed. |
| Right ventricular systolic pressure (RVSP) | Direct numeric variable of RVSP prioritized as the final result. Otherwise, free text analysis would select the number preceding “mmHg” as the RVSP. Erroneous values were manually corrected. |
| Diastolic dysfunction | Diastolic dysfunction determined using criteria in guidelines <sup>12</sup>. |
| Right ventricular variables | Free text analysis to identify right ventricular size and function |
| Export dataset | Dummy variables and raw variables dropped from the dataset. Dataset sorted by hospital number, and the earliest study date for each hospital number selected. Dataset was then exported. |

The data miner can save the search as a .xml file, allowing for reproducibility. In short, the data miner extracted all studies between 1 Jan 2010 and 31 Dec 2018 that were tagged as an echocardiogram. The specific variable names within the PACS can be found in this .xml, alongside a list of the final echocardiography variables these were used to generate. In total, 42,517 echocardiography studies were extracted.

#### De-identification

Data was de-identified in the following way

1. A random 10 character identifier (the “Anonymous ID”) was generated for each unique National Health Index (NHI) number. Each de-identified ID was linked to an NHI number in a lookup table.
2. Using the lookup table, NHIs in the main dataset were replaced with the Anonymous ID.
3. The lookup table was then saved and stored securely in accordance with the study protocol, separate to the de-identified data that were used for analysis.

### Echocardiography report variable cleaning

The data miner was unable to extract all studies into a single .csv file, due to computational power constraints. Hence, extracted studies were stitched together into a single dataset. All data cleaning and linkage was performed using R version 3.6.3 ^9^ using RStudio version 2021.09.01 ^10^ and the R packages “tidyverse” ^11^. The NA designation in variable refers to Not Available and labels missing data.

The order of data cleaning is summarised in **Table 1**.

#### Numeric variables

Numeric variables were all handled in a similar fashion: impossible or extreme values were recoded as NA, and the variable was otherwise kept the same. The following variables were handled in a unique fashion.

##### Body mass index (BMI)

BMI was calculated from height and weight (rather than being in the report itself). BMI was calculated as the participant’s weight in kilograms divided by their height (in metres) squared. Visual inspection of extreme BMI values revealed obvious instances where height and weight were transposed, and these were manually corrected (48 entries). A BMI ≥70kg/m^2^ or ≤14 kg/m^2^ was considered too extreme, and was recoded as NA.

##### Heart rate

Heart rate was often presented as a range of values and was recoded to be the mean of the two ends of the range. For instance, a heart rate of “120-180” was recoded as 150 beats per minute (bpm). Heart rates over 200bpm were excluded.

##### Right ventricular systolic pressure (RVSP)

For studies where the RVSP numeric variable was entered, this value was used directly. Otherwise, the free text of the report was analysed, and the numerals preceding a “mmHg” phrase were extracted. These were then coded as the RVSP. Entries were visually checked to ensure the value being coded was correct.

##### Removal of impossible values

For some variables, a negative value was not possible – for instance, LV posterior wall thickness cannot be zero or less. Therefore, for these variables, a value ≤0 was recoded as NA.

#### Categorical variables

Categorical variables were all handled in a similar fashion. The echocardiography report could contain either a true categorical variable (e.g. LV systolic function) or a free text section (e.g. left ventricle). In cases where the former was available, it was the preferred input. If not, then the free text section was analysed.

Free text is the major limitation to successful data analysis from a clinical database. However, frequently used phrases tend to become established in any echocardiography laboratory. Based on an initial set of phrases used commonly in local reports, a set of phrases were iteratively developed to detect free text entries that determined which level a categorical variable should fall into. An example of this process, using “aortic stenosis” severity, is presented in **Figure 1**. This code was developed iteratively – that is, studies would be classified by the code, and would be manually checked to see if there were any obvious misclassification errors. If there were errors (such as typographic errors or alternative ways of reporting), these were corrected by editing the R code. Some examples of different phrases are given in Table 2.

**Table 2:** Examples of phrases used to describe left ventricular function

| Phrase | Classification |
| --- | --- |
| systolic function appears within normal limits | Normal |
| LV function appears mildly and globally impaired | Mildly impaired |
| Systolic function is difficult to assess but visually appears mild-moderately impaired | Mild to moderately impaired |
| moderate decrease in systolic function | Moderately impaired |

**Figure 1:**
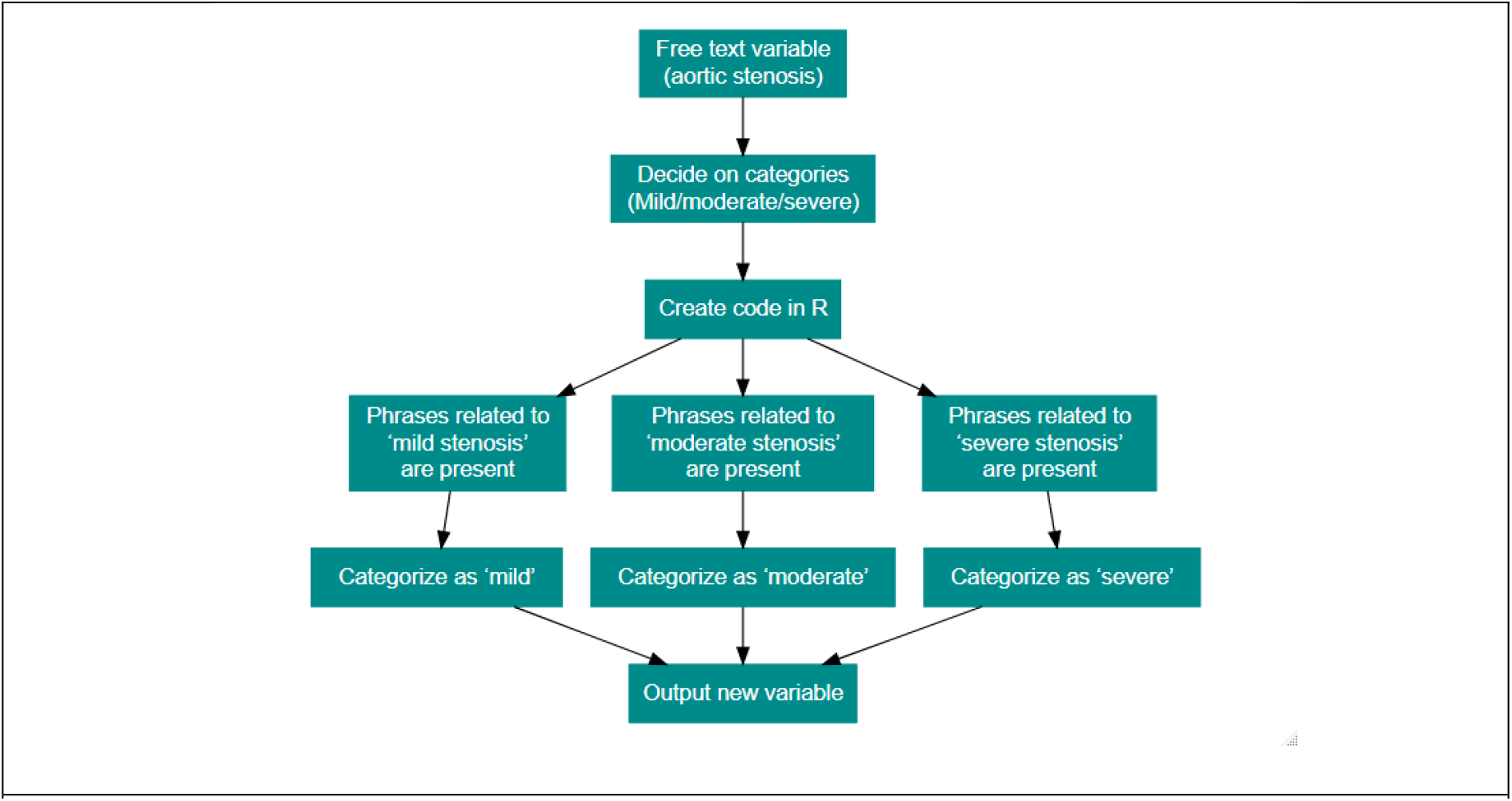
Categorization of free text entries into categorical variables

##### Categorization phrases

A selection of methods for certain variables are described below, because they were handled in a unique way or serve as a useful description for a method that was repeated for other variables.

##### Aortic stenosis

For full details of character terms in each category, please see the earlier linked R code. In brief, the following process was applied:

1. Whenever the word “thickened” or “sclerosis” is mentioned, “sclerosis” was reported as the CAVD severity
2. If any terms related to valve replacement (e.g. AVR, prosthesis, valve replacement) are mentioned, then the valve was assumed to be exogenous and was coded as an AVR
3. Each free text entry was then searched for terms relating to CAVD severity. If a term denoting CAVD severity was reported, then it was coded as that.

##### Aortic regurgitation

Aortic regurgitation (AR) was extracted in a similar fashion to that of aortic stenosis. The only key difference is that additional “catch-all” string searches were included – for instance, if the word “moderate” and “regurgitation” appeared in the same sentence together, the entry would be categorised as “moderate regurgitation”. Importantly, this function does not misclassify AR due to the order in which the terms are evaluated (see R code). For example, one could conceive that if the sentence described “mild regurgitation” and “moderate stenosis”, that the catch-all would interpret this is moderate regurgitation. This is not the case as the phrase “mild regurgitation” would have already been categorised.

##### Aortic valve morphology

Aortic valve structure typically falls within one of two categories – bicuspid or tricuspid. A very small portion of patients may have some other very rare morphology, and these studies were excluded due to very low numbers that would have likely limited patient anonymity. Entries containing the word “bicuspid” were categorised as “bicuspid”. Remaining entries containing the word “tricuspid”, stating the valve was normal, or not stating the valve structure, were classified as “tricuspid”. This was done as if the valve structure is not clearly stated, the treating clinician would believe it to be tricuspid. A bicuspid valve is a clinically significant finding and is very unlikely to not be reported, if identified on the echocardiogram.

##### Diastolic dysfunction

We utilised the ASE/EACVI guidelines to establish if participants had diastolic dysfunction ^12^. The decision tree is found in Figure 8 of these guidelines, and a “normal LVEF” for the two arms of the decision tree was defined as >50%.

##### Cardiac rhythm

Two separate variables were generated from the cardiac rhythm field in the report: a prioritised cardiac rhythm, and the presence of bundle branch block (BBB). BBB was categorised as left BBB, right BBB, BBB (unspecified), or no BBB.

Prioritized cardiac rhythm could have the following values:

‐ Artificially paced
‐ Supraventricular tachycardia (SVT)
‐ Atrial flutter
‐ Atrial fibrillation
‐ Heart block
‐ Sinus rhythm
‐ Other
‐ Uncertain

##### Left atrial size

Where possible, as it was the clinically used value, the free text interpretation of the LA size was preferred. However, at times, only the measurement was available without a qualitative size given. In these cases, LA size was defined according to international guidelines ^13^.

##### Remaining variables

All remaining free text variables (except for indications) were categorised in the same way: using the detection of phrases to place a study into the correct level.

##### Indications

A binary true/false variable was created for each indication category, and all participants were categorised as either fitting that indication (true) or not (false). A participant could have multiple indications. This level of detail was recorded to allow for more specific sub-analyses as well as the ability to collapse the categories down if simpler analysis was preferred. The full list of indication categories is reported in the supplement (supplementary table 2).

#### Data validation

Given the iterative nature of the data cleaning, a data validation set was essential. A random set of 100 studies were selected, and the R code output was compared to the final echocardiography report uploaded on the participant’s electronic health record. A Cohen’s kappa was calculated for each variable in the dataset (all numerical variables matched). In all, this process was repeated three times, with the final validation set reported here. This is because discrepancies were identified in the first two sets, and issues were subsequently corrected. The assessment of indication categories was derived by calculating the Cohen’s kappa for *each indication category*. The average Cohen’s kappa across all indications is presented in **Table 3**.

**Table 3:** Final validation set of data miner output ^a^ Mean value of all Kappa statistics for each indication category LV: left ventricule. LA: left atrium. RV: right ventricle.

| Variable | Proportion correctly matching clinical echocardiography report (%) | Cohens Kappa (k) |
| --- | --- | --- |
| Aortic valve stenosis | 97 | 0.95 |
| Aortic valve regurgitation | 100 | 1.00 |
| Aortic valve structure | 98 | 0.96 |
| Mitral valve stenosis | 99 | 0.89 |
| Mitral annular calcification | 99 | 0.94 |
| Mitral regurgitation | 95 | 0.86 |
| Tricuspid regurgitation | 99 | 0.99 |
| Pulmonary stenosis | 100 | 1.00 |
| Pulmonary regurgitation | 98 | 0.96 |
| LV size | 92 | 0.74 |
| LV function | 93 | 0.89 |
| LA size | 100 | 1.00 |
| RV size | 97 | 0.89 |
| RV function | 97 | 0.91 |
| Sex | 98 | 0.96 |
| Age | 100 | 0.98 |
| Indications | 95 | 0.88<sup>a</sup> |
| Cardiac rhythm | 99 | 0.98 |
| Bundle branch block | 100 | 1.00 |

## Results

## Discussion

This study aimed to develop a dataset linking clinically used echocardiographic data to outcomes in New Zealand. We aim to use this data to investigate the differences in survival and natural history of various conditions. We demonstrate excellent validity of our dataset, by comparing it against the clinically used electronic health record.

As mentioned in the background, recent work internationally has resulted in the use of several echocardiography databases to examine conditions of the cardiovascular system. The NEDA study, alongside databases from Israel and the United States of America have examined outcomes in conditions such as moderate aortic stenosis, pulmonary hypertension, and mitral, tricuspid, and aortic regurgitation^3-5, 7, 8^. We aim to further add to this literature using this generated dataset.

Our dataset has several strengths. Firstly, validation of the output by comparison with the electronic health record means that we can be confident that the variables represented are accurate to the clinical information used by healthcare providers. Secondly, we were able to link patients to granular outcome data, including surgeries, allowing for the potential of more in-depth analyses. Finally, because our data cleaning uses free, open-source software, our methods can inform future techniques for other researchers who may be considering a similar type of analysis.

Our intention is to use this data to examine the natural history of early calcific aortic valve disease, and the epidemiology of other forms of valvular abnormalities in a New Zealand population. However, our approach could be used by other clinical or research teams on the databases powering their own PACS systems, in order to answer clinical research questions or perform quality improvement studies. The PACS system is more than a simple image repository: it contains great potential to answer clinically relevant research questions and thus improve our ultrasound practice. The challenge is accessing the data in an accurate and meaningful way.

## Data Availability

The data produced for this study are unlikely to be available due to ethical and legal constraints

