## Supplementary table for "Use of an ultrasound picture archiving and communication system (PACS) to answer research questions: description of data cleaning methods"

### Syngo valve methods supplementary material

#### Supplementary table 1 – list of extracted variables

**Demographics**

- Study date
- National Health Index number (transformed to Anonymous ID)
- Age
- Sex
- Prioritized ethnicity
- Indication

**Anthropometrics**

- Height
- Weight

**ECG related**

- Cardiac rhythm
- Heart rate

**Aortic valve**

- Aortic valve morphology
- Calcific aortic valve disease severity
- Aortic regurgitation
- Aortic valve Vmax
- Aortic valve mean pressure gradient
- Aortic valve VTI

**Mitral valve**

- Mitral valve prolapse
- Mitral stenosis
- Mitral annular calcification
- Mitral regurgitation
- Mitral E Vmax
- Mitral A Vmax
- Mitral e'
- Mitral deceleration time

**Tricuspid valve**

- Tricuspid regurgitation
- TR Vmax

**Pulmonary valve**

- Pulmonary stenosis
- Pulmonary regurgitation

**Left atrium**

- Left atrial size

**Right atrium**

- Right atrial size

**Left ventricle (LV)**

- LV internal diameter end-systole (LVIDs)
- LV internal diameter end-diastole (LVIDd)
- LV posterior wall at end-diastole (LVPWd)

- Interventricular septum at end-diastole (IVSd)
- LV size
- LV systolic function
- LV ejection fraction
- Diastolic dysfunction
- LVOT VTI
- LVOT diameter end systole

**Right ventricle**

- RV size
- RV systolic function
- RV systolic pressure

**Aorta**

- Ascending aorta diameter

Aortic root diameter

#### Supplementary table 2 - indication categories

- ACS or MI (including coronary artery dissection)
- Aortic valve disease (including AS or AR)
- Congenital heart disease, but not bicuspid aortic valve
- Aorta (e.g. ascending aortic aneurysm, aortic coarctation)
- Atrial fibrillation (including atrial flutter)
- Malignant arrhythmia (cardiac arrest, VT, or VF)
- Other arrhythmia
- Pre-surgery (if explicitly stated the indication was pre-surgical)
- Post-surgery (any surgical indication was assumed to be post-surgery)
- Mitral valve disease
- Other valve disease
- LV function
- RV function
- Hypertrophic cardiomyopathy
- Dilated cardiomyopathy
- Other cardiomyopathy
- Chemotherapy/radiation therapy/oncology
- Chest pain
- Shortness of breath
- Heart failure
- Chronic coronary disease
- Murmur
- Pericardial disease
- Pulmonary hypertension
- Stroke or embolism (including any focal neurological signs, transient ischaemic attacks, or investigations for the source of a thrombus)
- Syncope (including any light-headedness, dizziness, or “funny turns”)
- Renal disease

- Diabetes mellitus
- Palpitations
- Heart infection (e.g. endocarditis or myocarditis)
- Rheumatic fever
- Hypertension otherwise unspecified
- Tachycardia
- Bradycardia
- Effusion or tamponade
- Implanted device
- Cardiomegaly
- Abnormal ECG
- Raised troponins
- Pulmonary oedema
- Non pulmonary oedema
- Other
